# The Hospital Murals Evaluation: A mixed-methods evaluation of the impact of murals on patients, healthcare workers, and visitors

**DOI:** 10.64898/2026.04.09.26350479

**Authors:** Nisha Sajnani, Marcel W. Foster, Yewande Oshodi, Kehinde Aniyat Sodimu, Mojca Kolnik, Marko Pokorn, Nicola Simpson, Tim Shaw, Simon Willmoth, Monica Marino, Larissa Trinder, Ebisan Akisanya, Ebere Onyekachi, Elisabeth Bahr, Victoria Blanchette, Research Assistant, Ebony Bolt, Qiaowa Gong, Haley Moyse Fenning, Deborah Olaitan Komolafe, Raphiel Murden, Nengi Omuku, Carl Rowe, Cris Sanhueza, Tim Steer, MacKenzie D. Trupp, Niamh White, Yaning Wu

## Abstract

**Objective:** This study investigated how hospital murals influence the experiences of patients, healthcare staff, and visitors across four sites.

**Background:** Evidence shows that visual art in healthcare settings can improve well-being but few studies focus specifically on murals or compare their effects across cultural contexts. The Hospital Murals Evaluation project addresses this gap through an investigation of murals in hospitals in Nigeria, Slovenia, the United Kingdom, and the United States.

**Methods:** Using a mixed methods cross-sectional design, the study integrated surveys, interviews, and participatory photography. A total of 525 unique responses were collected from 229 patients (131 adult, 98 pediatric), 245 staff, 49 visitors, and 2 undisclosed.

**Results:** Interviews across all three participant groups (n=115) revealed themes of positive affects, perception of care, as well as stress or indifference. Surveys (n=327) showed moderate positive correlations between mural viewing and positive emotions among patients, and between mural exposure and well-being, positive emotions, social connection, and workplace belonging among staff in the UK and USA, with null findings for staff in Nigeria and Slovenia. Participatory photography (n=83) illustrated how murals conveyed comfort, though abstract or poorly placed murals sometimes evoked discomfort. Meta-inferences across the methods indicate that viewing murals were associated with positive emotions for patients and did not induce negative emotions for staff or visitors.

**Conclusion:** Murals act as health-promoting infrastructure that can enhance well-being, foster positive experiences, and signal intentions of care. The findings highlight the need for culturally attuned designs to create healthcare environments that nurture well-being.

## Background

Hospitals are traditionally associated with clinical efficiency and medical interventions, yet for many they represent spaces of discomfort and stress during vulnerable life periods. In recent decades, research has increasingly highlighted the profound influence of architectural design, music, and visual art in shaping care environments and optimizing health outcomes (Laursen et al., 2014; Trupp et al., 2025; Viola et al., 2023). Within these environments, receptive engagement with visual art is increasingly recognized as a significant health resource, with growing evidence suggesting benefits for psychological well-being and belonging (Foster et al., 2025).

A substantial body of research demonstrates that art viewing, whether as part of everyday life, structured interventions, or hospital-based encounters, can support thriving and well-being (Cotter et al., 2022; Schall et al., 2018; Totterdell & Poerio, 2021). Even brief exposure, such as a single museum visit or engagement with one artwork, has been shown to reduce stress, negative mood, and anxiety, with similar benefits reported in hospital galleries (Fekete et al., 2022; Ho et al., 2015; Trupp et al., 2023).

Within healthcare environments specifically, visual arts, including paintings, murals, sculpture, digital projections, and interactive installations, have been integrated to foster healing, enhance resilience, and strengthen social connection (Foster et al., 2025). Reported outcomes include reduced heart rates among pediatric patients (Pearson, 2019), reductions in anxiety and stress symptoms (Gore, 2022; Lone, 2021; McCabe, 2013), decreased work-related stress among staff (Huet & Holttum, 2016), and improved perceptions of care quality and well-being among visitors (Abulawi, 2023; Monti, 2012).

A recent scoping review synthesizing 68 publications across 20 global locations concluded that visual art in healthcare settings can improve well-being, foster belonging, and enhance visitor experience, while also revealing important gaps in the literature (Foster et al., 2025). The majority of studies focused on patients, with fewer examining the experiences of staff and visitors. Notably, 28% of included studies (19 of 68) focused on murals, underscoring their prevalence. Murals, as large-scale and highly visible artworks, are particularly impactful in that they are accessible to multiple groups simultaneously, often carry cultural and symbolic meaning, and may influence not only individual health outcomes but also organizational culture.

The scoping review also highlighted that the effects of art interventions are not uniform and that cultural adaptation is essential for relevance, accessibility, and sustainability. Arts and health practices are deeply embedded in local values, beliefs, and aesthetic traditions, meaning that an approach effective in one setting may not yield the same outcomes elsewhere (Smith, Warran, & Fietje, 2025). An evaluability assessment conducted in preparation for this study reinforced this point, suggesting that the impact of hospital murals depends heavily on the cultural resonance of their imagery (Foster & Sajnani, et al., 2024).

This convergence of evidence reveals a critical gap: while research demonstrates that receptive engagement with visual art benefits health, and while murals are among the most prevalent art forms in healthcare settings, relatively few studies have rigorously examined their specific role or explored how their impacts vary across cultural and organizational contexts.

### Purpose of this Study

Building on these identified gaps, the Hospital Murals Evaluation (HoME) was designed as a cross-cultural, multi-site study to investigate the effects of murals in healthcare environments on the well-being of patients, healthcare staff, and visitors. As the largest study of its kind, HoME spans four countries, Nigeria, Slovenia, the United Kingdom, and the United States, each with distinct cultural, institutional, and clinical contexts (Figure 1).

**Figure 1.**
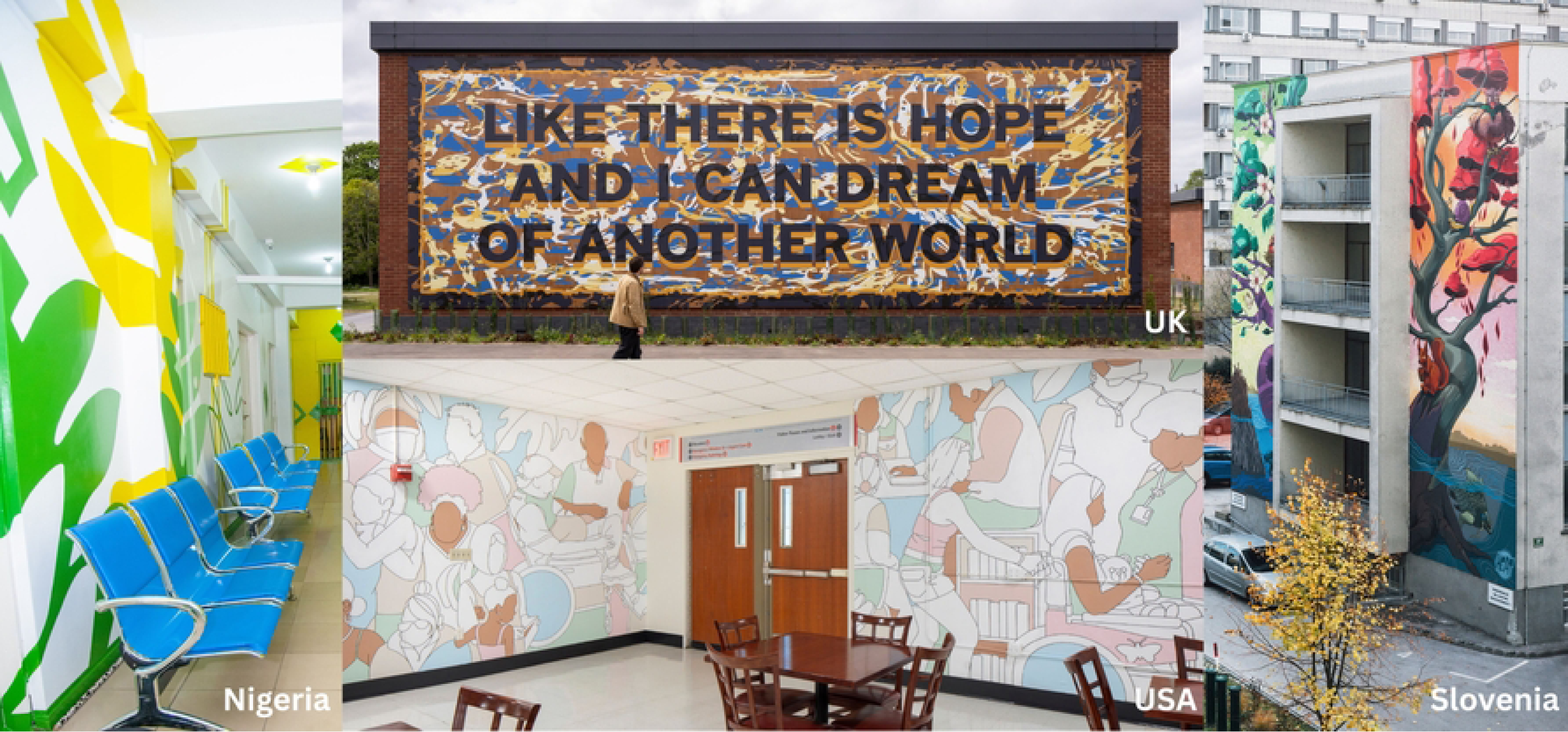

By including diverse participant groups across these international sites, this formative evaluation explores both consistent and context-specific impacts, offering critical insight into how murals can function as a culturally adaptive and accessible health resource (Tebes et al., 2015).

## Methods

This study employed a mixed-methods cross-sectional design to capture a comprehensive snapshot of how hospital murals impact patients, staff, and visitors across four culturally distinct healthcare settings. (Table 1) This study followed a culturally competent evaluation framework (CDC, 2014; Thomas & Campbell, 2020).

### Ethical Considerations

This research received ethics approvals from all participating sites (Nigeria: Lagos University Teaching Hospital: 6921; Slovenia: Ljubljana Children’s Hospital: 0120-372/2024-2711-6; UK: Norwich University of the Arts: 1-SW-2425; USA: NYC Health + Hospitals: 24-08-540-11HHC), as well as New York University (IRB8202). Participants Data collection occurred across the four sites from April - July, 2025 by staff, students, and interns affiliated with the site hospital and/or local research partner institution. All interviewees at all sites documented consent to record and use materials for the study and future publications. Ethics approvals included a waiver of informed consent for surveys and the participatory photography, as no personally identifiable information (PII) was collected.

### Healthcare Settings & Mural Interventions

Four hospitals were chosen based on existing relationships, the presence of murals at the facility, and the availability of staff to support onsite data collection. Each had maintained a visual art and mural program for over a year and contained multiple murals, defined here as images on the wall or ceiling of the healthcare setting. (Table 1).

### Data Collection Instruments

Data collection consisted of demographic questions, a survey consisting of original mural related questions and validated scales, semi-structured interview guidelines, and photo protocol which were refined and pilot tested between August 2024 - January 2025 with key stakeholders. Furthermore, Norwich University of the Arts engaged lived-experience teams to provide insights related to the relevance and sensitivity of all questions and delivery mechanisms.

### Data Analysis

#### Thematic analysis of interviews

A team of qualitative reviewers (MWF, OK, CS, VB, YW) reviewed transcripts using abductive coding (Vila-Henninger et al., 2022), synthesizing deductive and inductive codes to identify reflexive themes (Braun & Clarke, 2006; Byrne, 2022). MWF analyzed data from all sites, and other reviewers were assigned to one site based on their geographic location/cultural experience and/or interest (i.e., Nigeria: OK; Slovenia: VB; UK: YW; USA: CS). Using grounded theory (Deterding & Waters, 2021; Glaser & Strauss, 1967), inductive codes were applied to transcripts as well as deductive codes (Buroway, 1998). The deductive codebook was developed using literature identified on the health effects of visual art in healthcare settings (Foster et al., 2025). Coding was structured using the rapid and rigorous qualitative data analysis (RADaR) approach (Watkins, 2017). (Figure 2)

**Figure 2.**
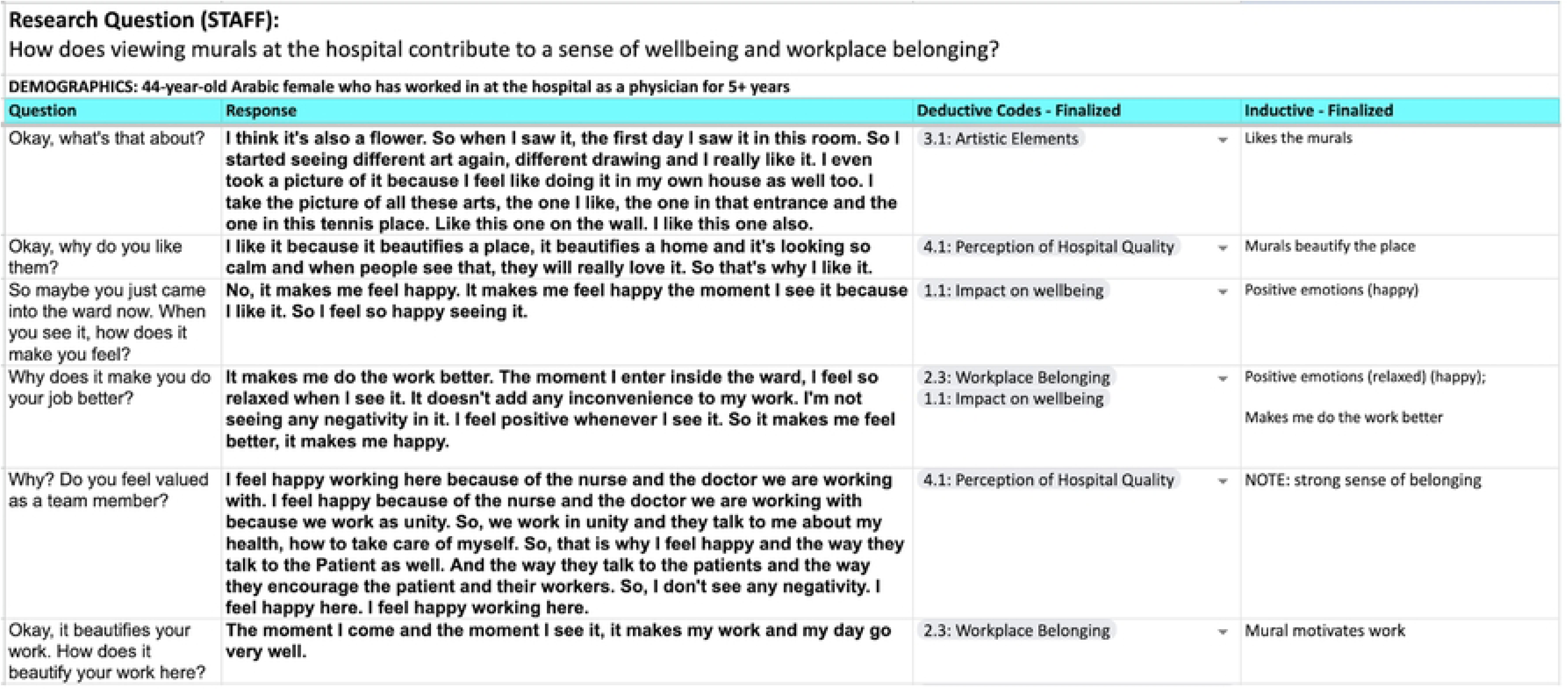

#### Quantitative analysis of surveys

All quantitative data analyses were conducted using R version 4.5.1 and simultaneously stratified by site and patient/staff status to avoid combining disparate populations. The two sections of the MES survey, as well as the validated scales, were examined for reliability using Chronbach’s alpha (Cronbach, 1951). Counts and percentages were calculated to describe demographic characteristics. We examined the distributions of MES and well-being scores via visual assessment of histograms and using the Shapiro-Wilks test of normality (Shapiro & Wilks, 1965). MES and well-being scores were described with means and standard deviations if they were symmetrically distributed. For those with non-symmetric distributions, we used medians and interquartile ranges. Similarly, either Pearson’s (symmetric distributions) or Spearman’s (non-symmetric distributions) correlation coefficients were employed to determine the correlation (Rodgers & Nicewander, 1988) between MES and each well-being score. Correlation p-values were Bonferroni-corrected (Dunn, 1961) within each site- and participant-type-specific population.

#### Textual and visual analysis of participatory photography contributions

The analysis of photos and the associated text relied on the Textual-Visual Thematic Analysis approach (TVTA) (Trombeta & Cox, 2022) and additionally leveraged RADaR (Watkins, 2017) to organize the findings.

#### Mixed methods analysis

Findings from the interviews, surveys, and participatory photography were analyzed at each site as well as across sites to identify points of convergence and meta-inferences through a joint display (Creswell & Plano Clark, 2017).

### Participants

A total of 525 unique responses were collected across three forms of data collection. (Table 2) Of these, 229 responses were from patients, of which 131 were adult patients and 98 were pediatric patients. 245 were staff, 49 were visitors, and 2 preferred not to disclose. All participants confirmed being exposed to one or more murals at their site and recruitment details, including specific incentives provided, are included in Appendix S1.

### Interviews

The 115 interview participants included 37 patients, 44 staff, and 34 visitors. (Appendix S2)

### Surveys

The 327 participants included 162 patients (Appendix S3 and S4) and 165 staff (Appendix S5).

### Participatory photography

Among the 83 participant contributions were contributions from 29 adult patients, 1 pediatric patient (USA), 36 staff members, 15 visitors, and 2 who preferred not to disclose. (Appendix S6).

## Results

### Qualitative Findings

#### Cross-cutting themes

For each population one to two quotes were selected to depict thematic nuances (Table 3).

##### Adult psychiatric patients (Nigeria & UK)

Adult psychiatric patients (n=18) reported themes of ‘Positive Affects’ as well as ‘Indifferent Affects.’ The patients also shared insights related to how the ‘Murals Reflect Intention of Care’ at the psychiatric inpatient settings (Table 4), as quoted below:

> “It makes you feel, when you look at the walls and us being here, it makes you feel like it’s a place that is built for patients to have a good experience. When you look at the paints and the murals, it gives a feeling that the intention of them was to help and not to cause madness**”** (Nigeria Patient 04).

##### Pediatric patients (Slovenia & USA)

Pediatric patients (n=19) reported six themes related to viewing murals at the hospital, including: ‘Positive Affects’ with insights related to how the murals were calming as well as providing a ‘Positive Distraction’, ‘Deep Reflections,’ and patients shared an ‘Increased Sense of Safety,’ as one patient shared that the murals “portray a safer environment” (USA Patient 01). Conversely, ‘Negative Affects’ and ‘Indifferent Affects’ were also raised by patients, with one patient expressing: “[The mural] is unpleasant, aggressive maybe. I mean if they’re for kids, they’re more appropriate. But when you’re a little bit older, they’re a bit stupid, let’s say, in the sense that they are pointless to you.” (Slovenia Patient 01). (Table 4)

##### Staff (all sites)

Staff interviews (n=44) included 30 clinical personnel and 14 non-clinical personnel. Participants shared insights that were categorized into a total 11 themes, which were organized into three prevalence categories, reflecting the number of sites that contributed to each theme. Five themes were identified across all four sites, which included: ‘Staff Observed Positive Impact on Patients,’ ‘Positive Affects’, and ‘Belonging,’ with a non-clinical staff member who shared “the murals make me feel good because each time I enter here to see them, they always make me feel at home” (Nigeria Staff 07). Similar to the patients, staff also expressed experiences of ‘Negative Affects’ and ‘Critiques of Wellness Initiatives’; with a clinical staff member sharing “We do need to get paid a little bit more to really take advantage… Otherwise, it’s just like all these things feel like band aids.” (USA Staff 09)

Additionally, three themes were identified across three of the sites, and these included: ‘Indifferent Affects’; ‘Insights on Co-Creation’; and ‘Social Connection.’ And three themes were identified across two sites, which included: ‘Positive Distraction’; ‘Murals Reflect Intention of Care’; and ‘Motivation to Provide High Quality Services’. (Table 4)

##### Visitors (all sites)

Visitors (n=34) shared insights that were analyzed into six total themes, which were organized into three prevalence categories, reflecting the number of sites that contributed to each theme. Two themes were identified across all four sites, which included: ‘Positive Affects’ and ‘Positive Distraction’, with a visitor sharing that the murals “[…] bring a bit of humanity into a clinical space”. (UK Visitor 04).

Additionally, three themes were found across three sites, including:‘Social Connection’; ‘Negative Affects’; and ‘Murals Reflect Intentions of Care.’ And two themes were identified at two sites, which were ‘Indifferent Affects’; as well as ‘Perceptions of Public vs. Private Hospitals’, with a visitor who shared “I was really impressed because I know [hospital name] to be a government facility, so for them to have thought of something like this impressed me.” (Nigeria Visitor 07). (Table 4)

### Quantitative Findings

#### Reliability and descriptive statistics

The original MES survey included high reliability across all participant types and sites (i.e., 0.72 - 0.95) whereas there was a wider range of reliability coefficients for the validated scales. (See Table 5) The MES survey indicates that most respondents tended to ‘Slightly Agree’ - ‘Agree’ related to questions about positive experiences in viewing the mural, with the exception of UK staff. (Figure 3; Table 6) Descriptive statistics for validated scales are included in Table 6.

**Figure 3.**
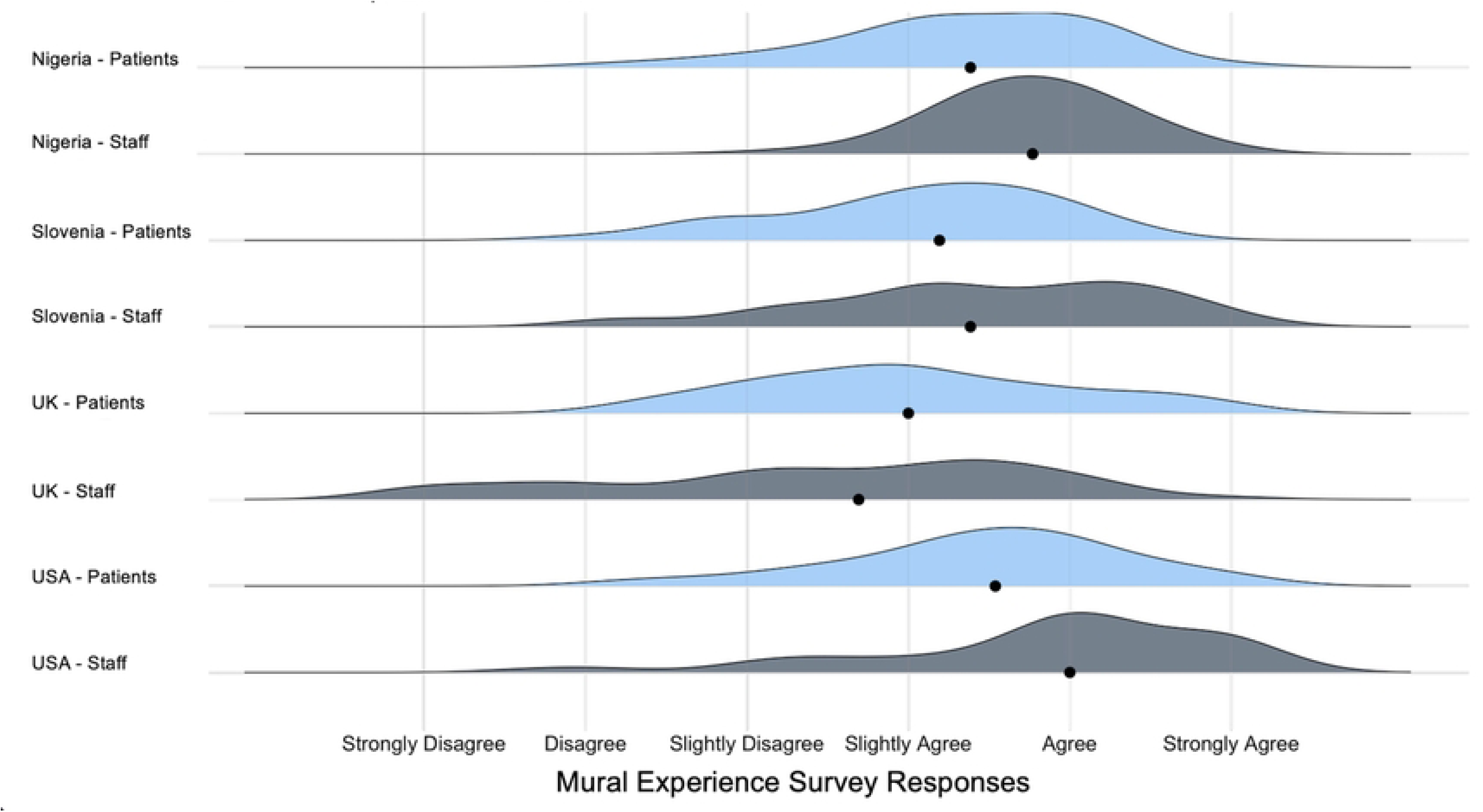

#### Cross-cutting and site-specific findings

Table 7 lists all correlations and Bonferroni corrected p-values. Adult patients in Nigeria (n=42) and the UK (n=42) had moderate positive correlations with viewing the murals and well-being, positive emotions, and social connection, but not negative emotions. Pediatric patients in Slovenia (n=36) and the USA (n=42) correlated viewing the murals with positive emotions, but data from patients in Slovenia did not show any other correlations. In the USA, correlations of viewing the murals as well as well-being and social connection were found; with both sites reporting no correlation with negative emotions. There were no statistically significant correlations between viewing the murals and well-being scales among staff in Nigeria (n=52) and Slovenia (n=17). Conversely, staff in the UK (n=50) and USA (n=46) reported significant correlations for viewing the murals and well-being, positive emotions, social connection, workplace belonging; neither site was found to correlate mural viewing with negative emotions. (Figure 4)

**Figure 4.**
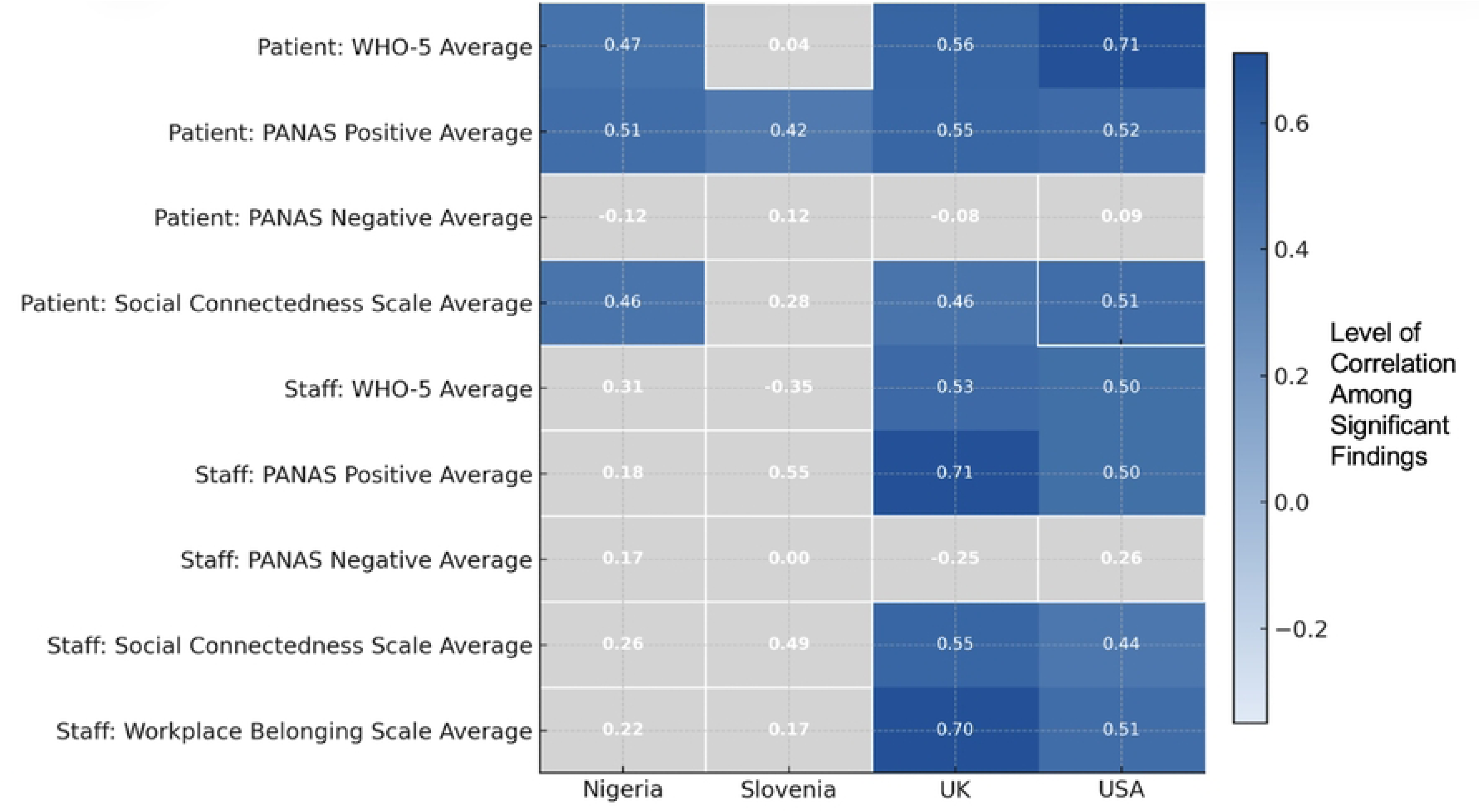

### Arts-Based Findings

#### Photo-textual analysis findings

In the ‘Reinforce’ category, there were 5 instances in the UK, 16 in the USA, and 6 in Nigeria. The ‘Complementary’ category comprised 10 instances in the UK, 16 in USA, and a higher count of 29 in Nigeria. ‘Contradictory’ codes were rare across all locations, occurring only once in the USA. Two responses were not able to be categorized. In aggregate, the total number of coded instances was 15 in the UK, 33 in USA, and 35 in Nigeria (n=83).

#### Cross-cutting themes

Using TVTA, we found the following themes across the sites. (Table 8)

##### Patients

Adult patients (n=29) participated across the three sites, with 10 in the UK, 6 in USA, and 13 in Nigeria. Adult patients were the largest participant group across all sites; only 1 pediatric patient participated, located at the USA site. Sites are noted as “cross-cutting” if reported by two or more sites.

Across sites, patients (n=30) most commonly described murals as evoking ‘Positive Affect,’ encompassing both aesthetic appreciation and/or positive emotional responses. Murals were seen as adding vibrancy, lifting spirits, and creating moments of comfort within the hospital. Patients also noted experiences of ‘Social Connection,’ as murals prompted conversations with others, or evoked memories of loved ones or home.

Participants linked murals, in some cases, to ‘Perceptions of Patient or Hospital Care.’ Patients also engaged in ‘Deep Reflection,’ interpreting murals through symbolic frames such as resilience, flourishing, love, safety, and peace. A full description of cross-site and single site themes are reported in Table 8. (Figure 5)

**Figure 5.**
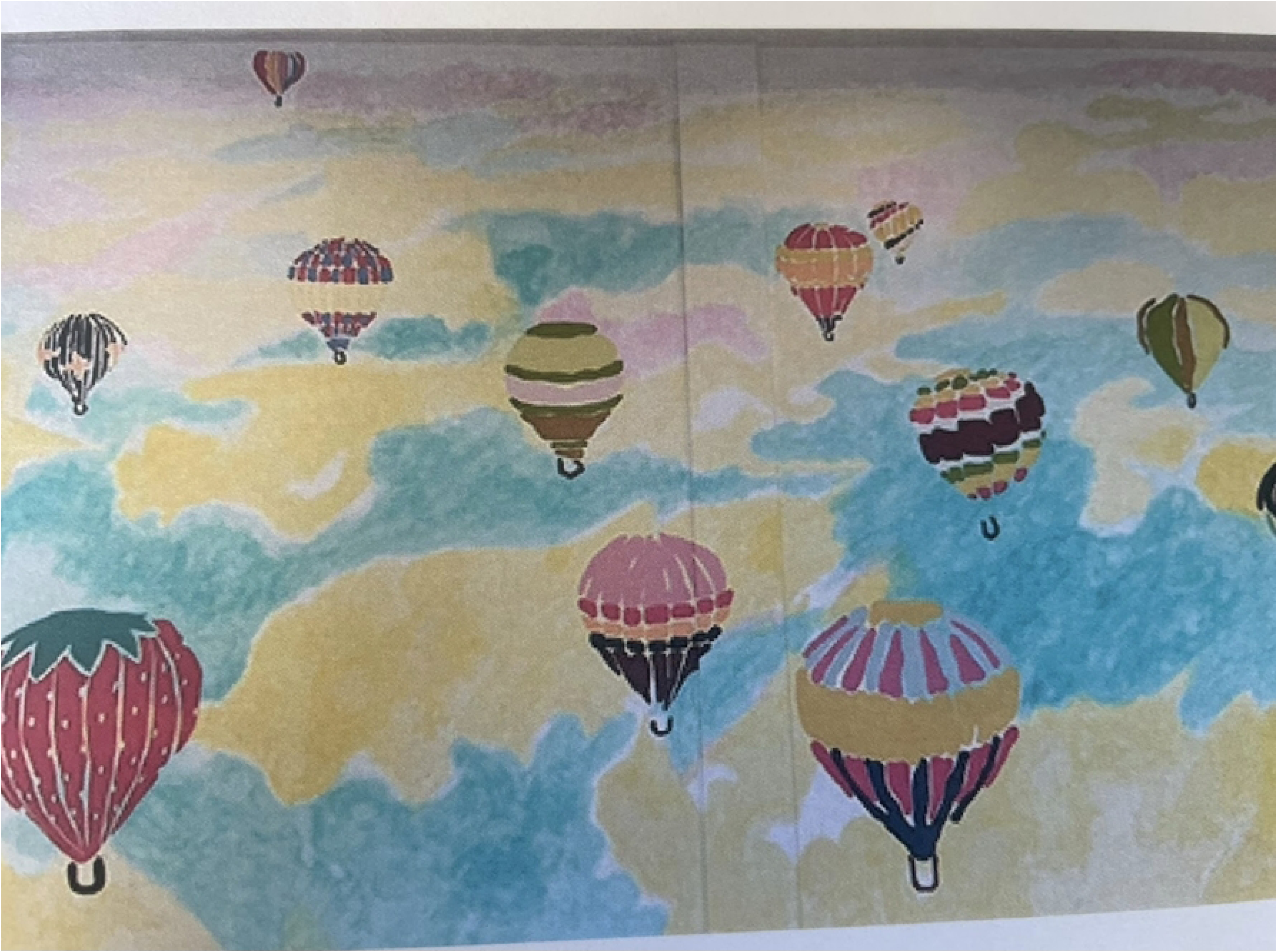
#“It’s abright, cheerful and therefore uplifting painting of balloons up in the air, dancing amidst the thermals, on a cloudy but sunny day. The multitude of balloons, 16 in total, all have distinctive liveries, play with colours. One of the balloons looks like a strawberry to hint of the time of the year, being the beginning of summer. All in all an uplifting, enjoyable visual” - PHT-UK-PAT-9

##### Staff

Staff participation (n=36) varied by site, with 2 staff members in the UK, 21 in USA, and 13 in Nigeria. Staff members emphasized murals as contributing to ‘Positive Affects,’ describing them as uplifting, inspiring, or aesthetically pleasing. Staff noted ‘Improvements to Workplace Perception,’ reporting that murals contributed to stronger positive identity within the hospital environment and served as a reminder of ‘Social Connection’. Some staff highlighted ‘Environmental Impact’, appreciating murals for adding vibrancy to institutional spaces. (Table 8) (Figure 6)

**Figure 6.**
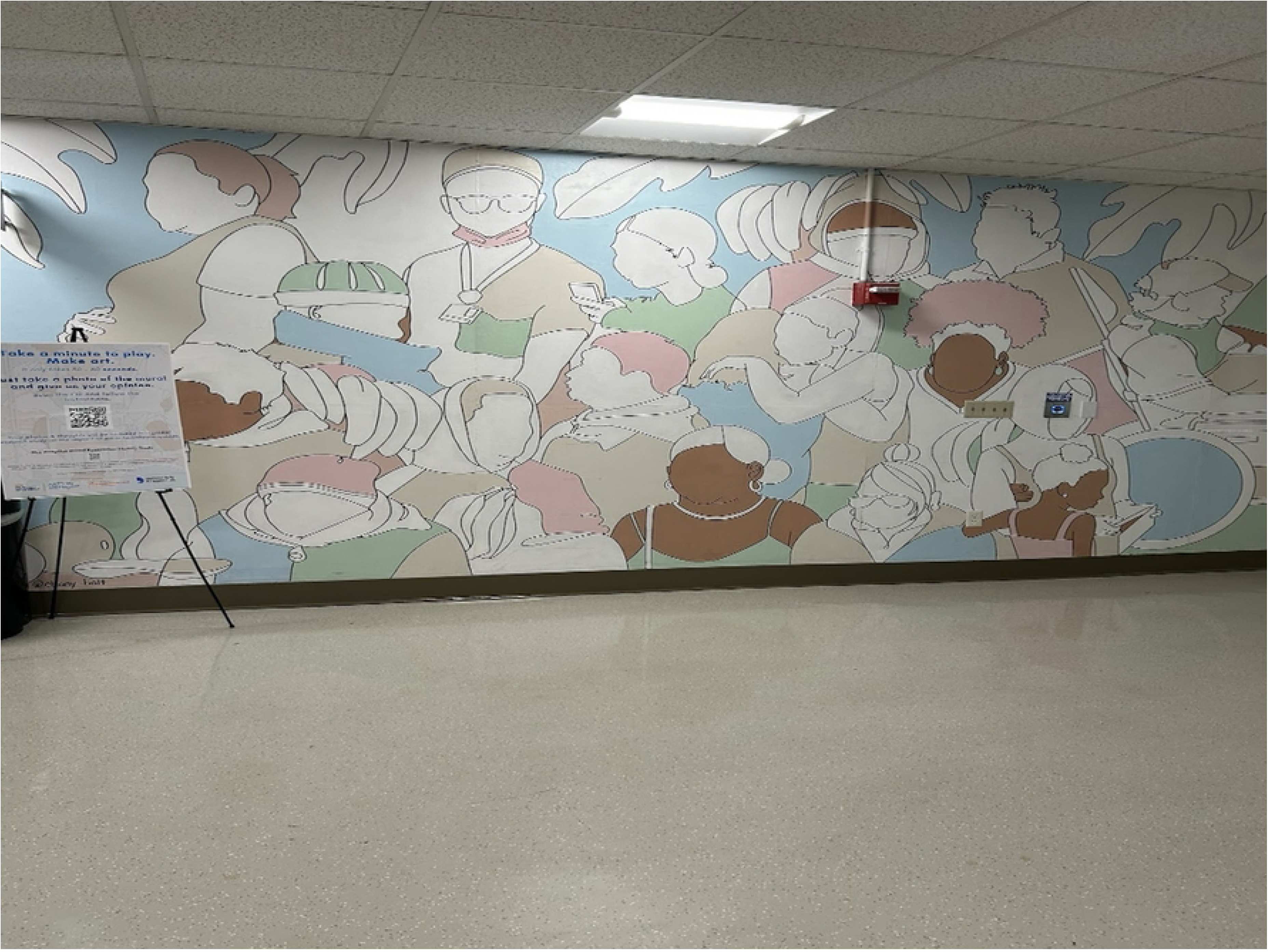
“I get a feel of coziness, unified, happy. It makes me want to eat in this space. It reminds me how hard the hospital work and how they become unified, together when it comes to patient care.” -PHT-USA-STF-16

##### Visitors

Visitor participation (n=15) included two from the UK, five from USA, and eight from Nigeria. Visitors framed murals as shaping their ‘Positive Affect’ through aesthetic enjoyment and inspiration Murals also facilitated ‘Social Connection’, acting as conversation starters. (Table 8) (Figure 7)

**Figure 7.**
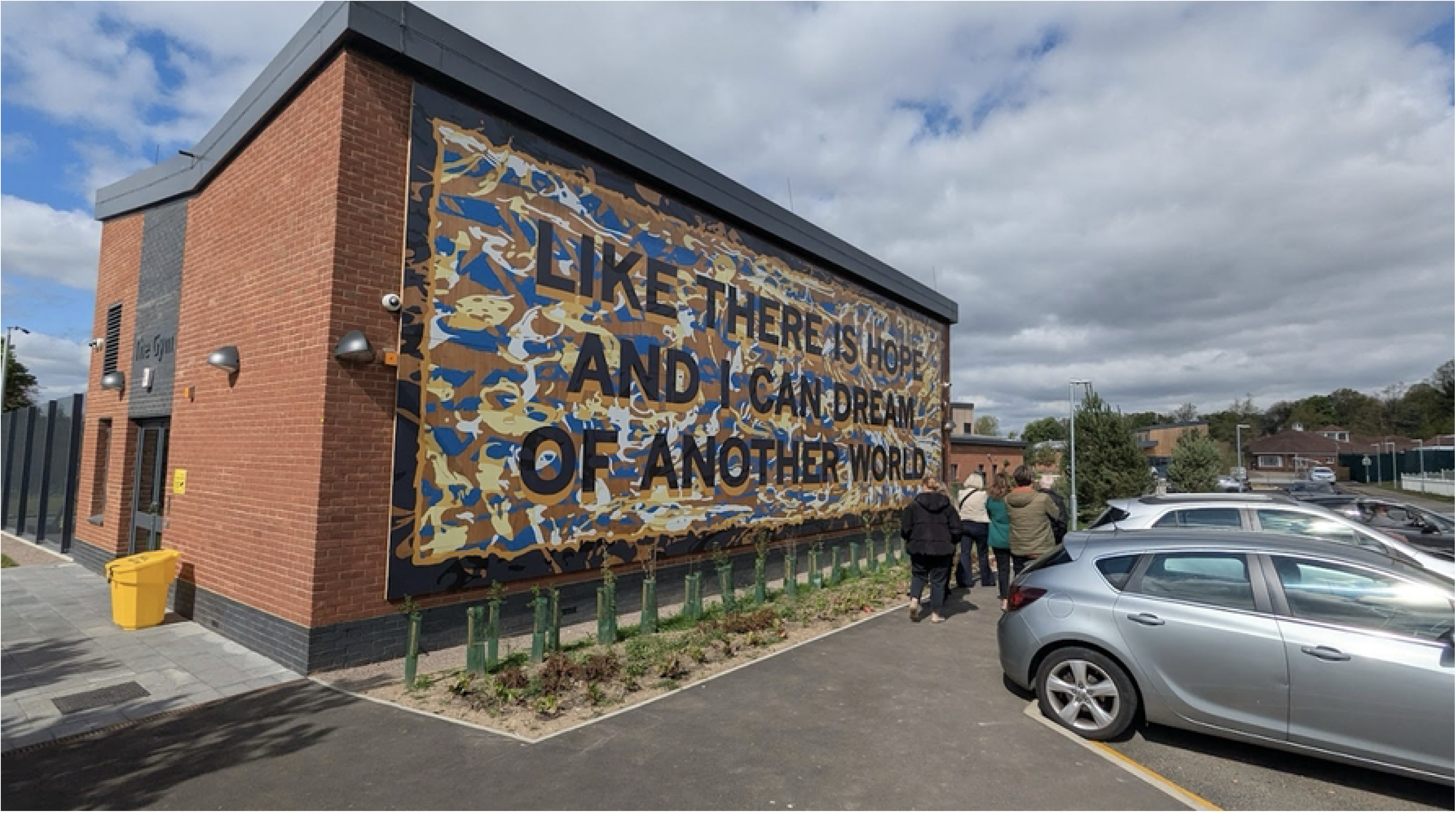
“The size and scale, colours and meaningful words. I think it represents something bold and you cannot ignore. I think it sparks discussion.” - PHT-UK-VIS-01

##### Participants who did not disclose role

There were participants who did not disclose their role (n=2) from Nigeria and the UK, and both were males. The themes identified for them related to biophilic design, deep reflection and positive affect. (Figure 8)

**Figure 8.**
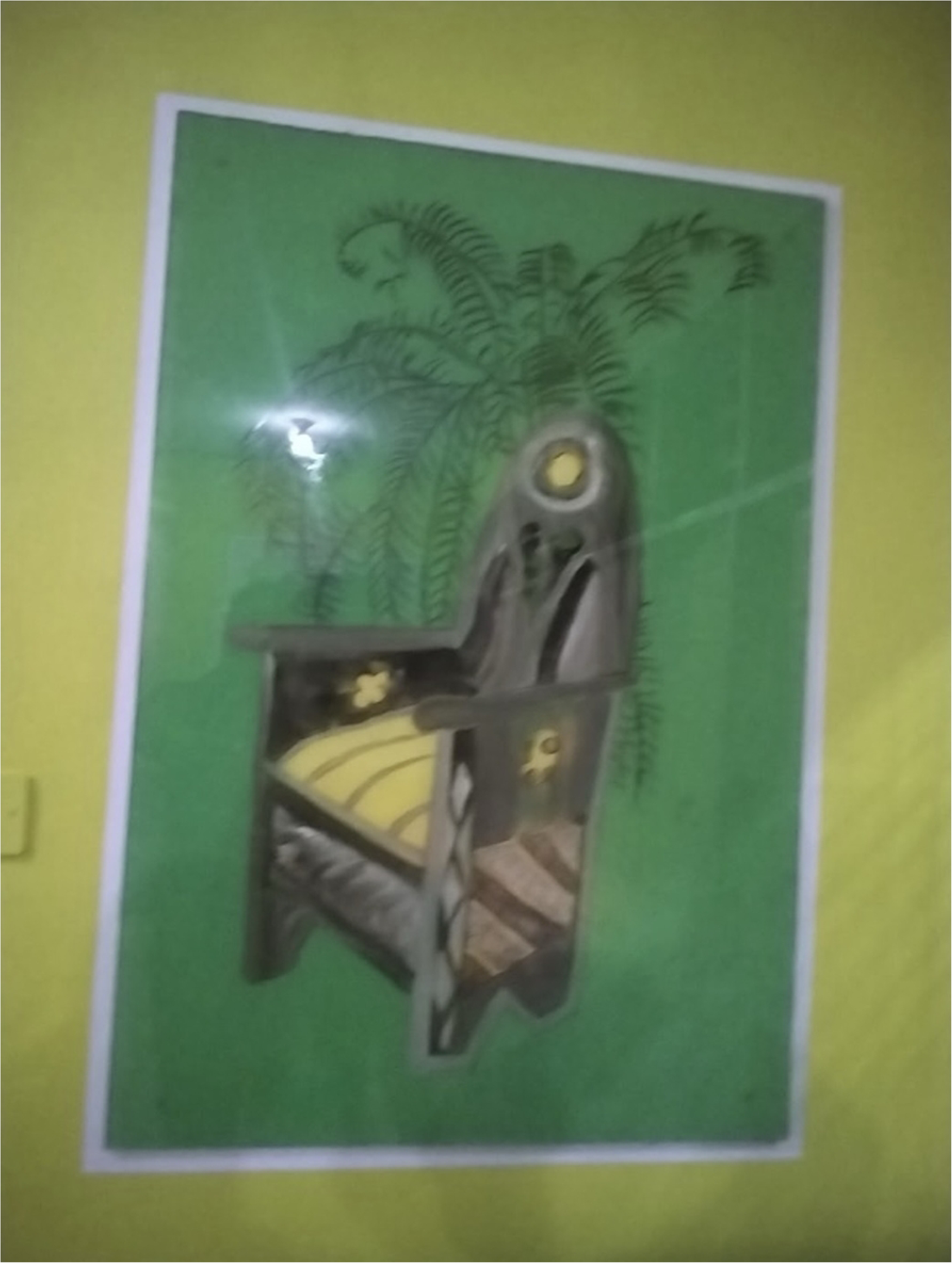
“Depends on my state of mind sometime it signifies that the fruit born by the palm tree are to be exploited since being a hospital ward, majestic chairs doesn’t tally with the ambience. But for a manic patients he or she may be excited that the throne belongs to him and he is a king.” - PHT-NIGERIA-NOT-1

### Mixed Methods Analysis

#### Cross-Cutting Findings

##### Patients

Adult patients (i.e., Nigeria and UK) were found to associate feelings of well-being as well as positive emotions to viewing the murals. Pediatric patients (i.e., Slovenia and USA) were found to associate positive emotions related to viewing the murals, with patients in Slovenia reporting this in survey responses and interviews, and American patients additionally contributing photos that align to this construct. Table 28 includes findings from both populations. Site-specific findings.

##### Staff

Surveys and interviews from Nigeria’s staff indicated that the murals may not be related with well-being, social connection, positive emotions, workplace belonging, and negative emotions. Interview themes aligned with a staff member asserting that increased consultation would be preferred to develop more relevant image content, connecting to the statistically insignificant finding with well-being. Contributions made by staff through the participatory photo methods were not able to align with these findings.

In Slovenia, surveys and thematic analysis of interviews indicated that staff did not associate viewing the murals with well-being, positive emotions, negative emotions, social connection, and/or workplace belonging. Interview themes supported these statistically insignificant findings related to negative and/or indifferent responses to the murals. In the UK, interview, survey, and participatory photo methods contributed to the insight that viewing murals may contribute to positive emotions (i.e., feelings of hope, peace, and aesthetic appreciation). Constructs of well-being, social connectedness, and workplace belonging also identified in quantitative and interview analyses, however, not supported by photo contributions, noting that only two staff members contributed to the participatory photo method. Finally, the null finding of negative emotions did not have clear inferences to the other methodological insights.

For staff in the US, the interview, survey, and participatory photo methods reinforced findings that viewing the murals were correlated with well-being (e.g., relaxation in the workplace), positive feelings (e.g., happiness), social connectedness, and workplace belonging. The null finding of negative emotions did not have clear inferences to the other methodological insights. (Table 9)

##### Visitors

As visitors did not partake in completing surveys, mixed methods analysis was analyzed using interview and participatory photography outcomes. Nigerian visitors reported that murals contributed to feelings of well-being (i.e., feelings of “peace and calming”) and also shared that the murals beautified the facilities. In the UK hospital, visitors shared insights that a mural that features text could likely help contribute to socially connecting to others, and particularly patients. And in the USA, visitors shared that murals beautified the facility, depicted community care, and reflected the area’s diversity. Table 9 includes findings from all sites.

## Discussion

### Murals as Health-Promoting Infrastructure

Findings from the HoME study indicate that murals should not be conceptualized as simply decorative features but as *health-promoting infrastructure*. Across four countries and diverse hospital settings, mural viewing was linked with positive emotions for pediatric and adult psychiatric patients, a finding that was supported across mixed methods. In particular, findings from the MES suggested that viewing murals was positively associated with constructs of well-being and aesthetic experiences. Interviews and arts-based methods indicated that participants consistently described murals as sources of comfort, calm, social connection, and workplace belonging for staff. These findings align with a growing body of evidence showing that visual art in healthcare environments can measurably influence well-being, reduce stress, and shape perceptions of care quality (Cotter et al., 2022; Laursen et al., 2014; Trupp et al., 2025). Participants frequently interpreted murals as markers of institutional investment and what we coded as an *‘*intention of care.*’*

### Perceptions of Murals Across Hospital Communities

Across all sites, participants engaged with hospital murals through distinct emotional and relational lenses shaped by their roles as patients, staff, or visitors. These differences in mural perception reveal how murals function simultaneously as personal, professional, and communal resources within healthcare environments.

Patients primarily discussed murals in *sensory and emotional* terms. Adults emphasized feelings of calm, comfort, and inspiration, particularly in response to biophilic imagery evoking renewal or escape. Pediatric patients described murals as playful and reassuring, helping transform hospitals into more welcoming spaces. Yet, some noted that abstract or age-inappropriate imagery diminished engagement, suggesting that developmental relevance is key to sustaining positive effects for pediatric patients.

Staff tended to view murals through an *organizational* lens. Many described them as contributing to workplace pride, morale, belonging, and as one clinical staff-member noted, “…a softness that art can bring to an environment for people’s well-being” (UK Staff 07). In Nigeria, the UK, and the USA, staff linked murals to improved emotional climate, yet some expressed ambivalence, noting that murals in corridors often went unnoticed or that certain designs felt mismatched with the clinical tone of the environment. In the USA, staff also raised themes of representation and diversity, identifying murals as opportunities for institutions to express inclusion and cultural respect.

Visitors highlighted *aesthetic and social* functions. They described murals as beautifying facilities, creating welcoming atmospheres, and acting as conversation starters between patients, staff, and families. In Nigeria, the UK, and the USA, murals that reflected local culture or community imagery inspired pride and connection, though some visitors found bright or complex designs overstimulating.

Together, these perspectives underscore that murals operate on multiple levels: as affective resources for patients, organizational signifiers for staff, and social anchors for visitors which link the hospitals to their communities. Recognizing these layered meanings reinforces the importance of participatory, context-sensitive approaches to mural design. Notably, across all sites, there was no statistically significant relationship between mural viewing and negative emotions in quantitative findings, consistent with the predominance of qualitative reports linking murals with positive feelings, distraction, and perceived care.

### Location and Content of Murals: Consistency and Cultural Nuance

While cross-cutting themes such as positive affect and distraction were evident across all sites, cultural and contextual differences emerged in both mural content and placement. Across sites, participants expressed distinct cultural and contextual preferences for mural content. In Nigeria, biophilic imagery (e.g. trees, flowers, and greenery) was viewed as a metaphor for abundance and community, while some visitors associated murals primarily with pediatric or psychiatric settings, reflecting limited exposure in non-specialized hospitals. In Nigerian and US sites, visitors associated commissioned visual artwork with privately funded hospitals, and were surprised to see murals created in public hospitals. In Slovenia, two trends based on age emerged: younger children preferred colorful and bold designs, while older youth found them childish. Staff mirrored this pattern as well, describing the imagery as both comforting and, at times, too juvenile for adolescents. Participants in the UK expressed divergent reactions to the inclusion of text: some found it meaningful and grounding, while others considered it distracting or even emotionally triggering. In the US, participants emphasized the importance of representation and diversity, viewing murals as reflections of inclusive institutional identity. Collectively, these findings highlight how cultural adaptation and contextual sensitivity are essential in designing effective arts-in-health interventions (Abulawi, 2023; Foster & Sajnani, et al., 2024; Smith, Warran, & Fietje, 2025).

In terms of location, staff and visitors indicated that placement shaped meaning and emotional impact. Murals in transitional spaces (e.g., corridors, atria) were at times overlooked, while those in waiting areas, break rooms, or patient wards invited reflection and comfort. However, alignment with context mattered: images of idyllic landscapes in a clinical area could evoke a sense of isolation from the depicted paradise, rather than create a sense of calm. In the UK, several staff commented on the importance of aligning content with the facility’s context (e.g., murals with vigorous content in a gym; murals with calming content in patient rooms). And in the USA, communal-space murals inspired appreciation among some and ambivalence among those who may have been visually overstimulated by the busy hospital environment. Overall, a mural’s placement, and its fit with the emotional and functional character of each space, proved central to its aesthetic, psychological, and social impact.

### Relational and Organizational Benefits

Beyond individual well-being, murals facilitated social connection among patients, families, staff, and visitors. In interviews, participants described murals as conversation starters, sources of shared reflection, and symbols of unity. Staff frequently associated murals with workplace belonging, echoing prior research linking visual art to improved emotional well-being and a stronger organizational culture (Butler et al., 2020; Huet & Holttum, 2016). Visitor perspectives added nuance, noting that text-based murals in particular could serve as tools for social connection, especially in relation to patients in longer-term care settings.

At the same time, there were notable variations in both single and multi-site findings in how participants experienced murals, highlighting the importance of cultural and organizational context in shaping impact. For hospital staff in Nigeria and Slovenia, no statistically significant correlations were found between viewing the murals and well-being, social connectedness, positive/negative emotions, or workplace belonging. However, interviews from both sites included personnel perspectives on the role of murals to promote positive affect, social connection, and workplace belonging. For staff in UK and USA hospitals, there were numerous meta-inferences supported by statistical correlations and the other methods related to how viewing the murals was associated with well-being, social connection, positive emotions, and workplace belonging. This could be, in part, because of the participatory processes involved in designing and/or working with completed murals at sites in the UK and USA.

As previously noted, these impacts are not uniform, underscoring the need for further inquiry into the relational, organizational, and cultural factors that determine whether, how, and for whom murals meaningfully contribute to both individual and collective health outcomes. Further, these findings suggest that there are complex, context-dependent factors (e.g. organizational culture, staff demographics and characteristics, or mural content) that can influence the effects that art in a hospital environment may have.

### Murals are Not a Panacea

Despite many benefits, the study also surfaced negative or indifferent responses. Some participants felt that abstract, text-based, or poorly placed murals were stress-inducing and/or emotionally unsettling. Staff critiques of wellness initiatives further suggested that visual interventions must be embedded in broader organizational strategies to address systemic concerns, such as staff shortages or inadequate compensation, to mitigate staff perceptions of superficiality. These critiques should be treated as *constructive feedback*, informing future mural initiatives that are responsive to both aesthetic and institutional realities.

### The Importance of Co-Creation and Meaningful Interaction

Across three sites, participants emphasized the value of participatory design in fostering authenticity and ownership. Interview findings indicated that involvement in mural creation enhanced appreciation and connection, aligning with evidence that co-creation strengthens perceived relevance and well-being of the hospital mural (Foster & Sajnani, et al., 2024; Luftglass & Rothschild, 2022). Participants also advocated for co-produced processes governing mural maintenance and removal, ensuring decisions reflect collective input rather than managerial discretion. Artists echoed this emphasis. As Nigerian artist Nengi Omuku noted, “Collaboration lies at the heart of this project… [including] engag[ing] directly with service users to understand what they envision for the space… and consult[ing] with providers to learn what elements they believe would be most therapeutic”. Similarly, Tim Steer from Hospital Rooms emphasized that “co-production is the foundation for creating something meaningful, relevant, and lasting”. Such insights re-affirm that murals achieve their fullest potential when grounded in genuine collaboration across organisational hierarchies.

### Sustaining the integration of arts in healthcare

The question of how murals, and the arts and arts therapies more broadly, are funded within hospitals carries important implications for sustainability and equity. Across international contexts, per the funding reported by partners involved in this project, mural projects have been largely supported through philanthropic donations, public–private partnerships, or artist-led initiatives rather than core hospital budgets. While such models have enabled the proliferation of creative projects, they can also lead to uneven distribution, dependence on external funding cycles, and limited continuity in maintenance or evaluation. Establishing a dedicated office, department, or role within healthcare systems to oversee arts integration offers a more sustainable and equitable approach to instill that patients, staff, and visitors can benefit from murals and art within hospital settings. Embedding arts leadership within health systems would move the arts from the margins of philanthropy into the core of healthcare planning, positioning them as integral components of patient-centered, culturally responsive, and organizationally coherent models of care.

### Implications for Policy and Practice

Taken together, findings suggest that murals represent *scalable* and culturally *adaptable* intervention that can be integrated into hospital planning and budgets as part of broader strategies to enhance healing environments, improve staff well-being, and foster patient and visitor satisfaction. As remarked in other studies of visual art in healthcare contexts, policymakers should prioritize participatory design approaches and iterative evaluation to ensure cultural and contextual relevance (Luftglass & Rothschild, 2022; NAHN, 2025). When institutionalized as infrastructure rather than optional decoration, murals can help embed equity, belonging, and dignity into the built environment of care. Acknowledging the arts as a central part of healthcare, rather than a peripheral or ‘nice-to-have’ element, is critical to transforming healthcare environments.

### Limitations and Future Directions

This study offers formative, cross-cultural insights into how murals are experienced in hospital environments, but several limitations must be acknowledged. First, the cross-sectional design captured a single moment in time, precluding causal inference or assessment of whether effects are sustained. Recruitment approaches varied across sites to align with local customs. While consistent with a culturally competent evaluation framework (CDC, 2014; Thomas & Campbell, 2020), this variation may have introduced inconsistencies in data quality and representativeness. Strategies such as digital tablets, QR codes, and paper surveys reflected cultural responsiveness but may also have affected the comparability of responses across sites. The variation in recruitment integrity across sites could have also played a role in lower survey completion rates, particularly among staff members in Slovenia. While sample sizes across methods were sufficient for exploratory analysis (between 83-327 participants), qualitative subsamples at each site (an average of 10 participants per informant type at each site) were relatively modest and may not have reached full thematic saturation. The statistical findings should also be interpreted cautiously, given the modest sample sizes, cultural variability across sites, and cross-sectional design. Additionally, our original MES survey was adapted from the WEMWBS and Aesthetic Experience validated scales to assess the extent to which individuals had diverse positive responses with the murals in the hospitals. Therefore, individuals were recalling one situation or multiple viewing sessions that they had with the murals, introducing potential recall bias. Furthermore, the development of the questions could have also indirectly measured a trait quite similar to aesthetic responsiveness or aesthetic sensitivity (Wanzer, et al., 2020), which is known to be related to well-being cross-sectionally and to well-being improvements via states of meaningfulness and pleasure during art viewing (Trupp et al., 2023).

While ‘potential for mural exposure’ was included as a demographic variable in our findings, stratification of the impacts of exposure to a mural by exposure duration was not possible due to logistical constraints (e.g., the mural at the USA site was finalized in December 2024). Participation in the ideation and creating of the murals occurred in Nigeria, UK, and the USA, however, inquiries about participants’ participation were limited only to the UK and USA. Measurement validity was also constrained: only the WHO-5 was available in a validated Slovenian translation, and other scales were newly translated for this study, potentially limiting sensitivity to cultural nuance; however, internal reliability was mostly strong across the scales for each site and population (i.e., mostly above 0.7), and mitigated some risks related to measurement validity. Although multiple check-ins with local staff were conducted to confirm accuracy of qualitative interpretations, not all cultural subtleties may have been identified. Translations of quotes from Slovenian to English, as well as the diversity of English dialects within and across each site, present limitations in the precision of thematic interpretations although this was somewhat mitigated through iterative consultation with site leaders. As murals were situated in different kinds of hospital spaces, including psychiatric wards, pediatric facilities, and public areas, comparisons across settings are tentative, and findings should not be generalized to all healthcare environments. Finally, participant compensation varied across the sites and this could have affected participation.

At the same time, a strength of the study was the development of the Mural Experience Survey (MES), which demonstrated high reliability across all four sites. More testing is likely required with larger sample sizes and with controlled samples. Future work should examine the scale’s convergent and discriminant validity, and potential factor structure. An additional strength of this study was its use of mixed methods, which allowed for qualitative insights to be supported by quantitative inferences and art-based data to support key findings. Indeed, the incorporation of arts-based methods for arts interventions provides critical nuances for analyses (Leavy, 2020). Given this study’s emphasis on thematic analysis and inclusion of inductive codes, future qualitative investigations on this topic could build on our findings to develop a content analysis guideline and report (Krippendorff, 2018).

Future research should build on these formative insights with longitudinal and experimental designs, including pre/post and randomized controlled trials, to more clearly assess validation between mural exposure and outcomes (e.g., experiences of stress, patient recovery times, and/or workplace belonging). Incorporating physiological and clinical measures, such as cortisol levels, heart rate variability, and blood pressure, alongside psychosocial scales and validated measures of mural experience, would strengthen evidence of health impact. Economic analyses are also needed to quantify the value of murals for healthcare systems, including potential contributions to patient satisfaction, staff retention, staff absence, and efficiency gains. Future studies would also benefit from controlling for environments within and/or across the healthcare facilities, to examine exclusively “public” settings (e.g., corridors or hallways) and/or private settings (e.g., patient rooms, staff breakrooms).

Further inquiry should also explore differences between co-created and non-participatory mural projects, examining whether process influences health and organizational outcomes. The emphasis among participants on representation and cultural relevance suggests that equity considerations are critical. Future studies should continue to employ mixed methods, integrating arts-based, qualitative, and statistical analyses, since qualitative insights add essential contextual nuance to quantitative findings. By advancing this evidence base, murals can be more firmly established as infrastructure for care, embedded into hospital planning, budgeting, and design standards worldwide.

## Conclusion

The HoME study provides the strongest cross-cultural evidence to date that murals function not merely as decoration but as health-promoting infrastructure within hospitals. Across four countries and diverse clinical contexts, patients, staff, and visitors consistently described murals as sources of comfort, positive distraction, and connection, while quantitative findings revealed positive associations of mural-experience with well-being, positive affect, social connectedness, and workplace belonging in certain sites. Importantly, cultural variations underscored the value of contextually relevant design and co-creation processes, demonstrating that murals can embody an authentic “intention of care.” These findings position murals as an adaptable intervention with the potential to not only improve individual and organizational outcomes in healthcare, but additionally to support and invigorate healthcare staff. By embedding murals into hospital planning, policy, and budgets, healthcare systems can advance both equity and quality of care, advancing the aim for environments of healing to also be environments of dignity, belonging, and resilience.

## Acknowledgements

We would like to thank Mary Peng and Varshini Odayar for their support on the evaluability assessment; Tolulope Akinmolayemi, Azeez Quadry, Akinbiyi Bolanle, Maja Buzuk, Lejla Kanuric, Carl Rowe, Nicola Simpson and Ryan Patrick for their assistance with data collection; Tessa Brinza and Yaashna Sharma for assistance with project management; and Jill Sonke for her insights. Authors are ordered alphabetically according to study site and last name.

## Data Availability Statement

Data and supplementary research materials may be accessed: https://shorturl.at/czmoH

